## Supplementary Material for "Assessing the Use of Prescription Drugs in Obese Respondents in the National Health and Nutrition Examination Survey"

### Supplemental Material

#### Use of NHANES Dataset

Table S1 lists the NHANES files, the NHANES variables, the associated questions, and how each variable is referred to in this paper.

**Table S1.** NHANES Variables

| **NHANES File** | **NHANES Variable Name** | **NHANES Question** | **Reference in the Paper** |
|---|---|---|---|
| Body Measures | BMXBMI | Body mass index | BMI |
| Demographic Variables and Weights | SEQN | Respondent sequence number | None |
| Demographic Variables and Weights | RIAGENDR | Gender | Sex |
| Demographic Variables and Weights | RIAAGEYR | Age in years at screening | Age |
| Demographic Variables and Weights | RIDRETH1 | Race/Hispanic Origin | Race |
| Demographic Variables and Weights | WTINIT2YR | Full sample 2 year interview weight | None |
| Demographic Variables and Weights | INDHHIN2 | Total household income | Annual household income |
| Dietary Supplement Use 30-Day - Individual Dietary Supplements | DSDCOUNT | Total # of Dietary Supplements Taken? (Includes all supplements and the antacids reported with supplements, but not antacids reported with medications.) | DS use |
| Health Insurance | HIQ011 | Covered by health insurance? | Health Insurance |
| Income | INDFMMPI | Family monthly poverty level index (Family monthly poverty level index, a ratio of monthly family income to the HHS poverty guidelines specific to family size.) | PIR |
| Prescription Medications | RXDUSE | Taken prescription medication, past month | RXD use |
| Prescription Medications | RXDCOUNT | Number of prescriptions medications taken | Number RXD |
| Prescription Medications - Drug Information | RXDDCN1A | Drug category name - CAT 1, LEV 1 | RXD category |

**Regression Analysis**

**Table S2.** Reported prescription drug use by demographic characteristics among obese and control group.

| Variable | Control | | | | Obese | | | |
|---|---|---|---|---|---|---|---|---|
| | Odds ratio | 95% Wald CL | | P value | Odds ratio | 95% Wald CL | | P value |
| **Gender** | | | | | | | | |
| Male | 0.572 | 0.538 | 0.607 | <.0001 | 0.552 | 0.508 | 0.6 | <.0001 |
| Female (reference) | 1 | | | | 1 | | | |
| **Age Group** | | | | | | | | |
| 18-24 | 0.258 | 0.21 | 0.316 | <.0001 | 0.298 | 0.224 | 0.395 | <.0001 |
| 25-34 | 0.364 | 0.298 | 0.444 | <.0001 | 0.367 | 0.282 | 0.476 | <.0001 |
| 35-44 | 0.426 | 0.35 | 0.52 | <.0001 | 0.39 | 0.303 | 0.503 | <.0001 |
| 45-54 | 0.531 | 0.436 | 0.646 | <.0001 | 0.569 | 0.443 | 0.73 | <.0001 |
| 55-64 | 0.77 | 0.631 | 0.94 | 0.0104 | 0.922 | 0.718 | 1.182 | 0.5207 |
| 65-74 | 0.853 | 0.727 | 1.001 | 0.0515 | 0.922 | 0.739 | 1.151 | 0.4742 |
| 75 over (reference) | 1 | | | | 1 | | | |
| **Race** | | | | | | | | |
| Mexican American | 0.587 | 0.519 | 0.664 | <.0001 | 0.658 | 0.565 | 0.765 | <.0001 |
| Other Hispanic | 0.762 | 0.659 | 0.881 | 0.0002 | 0.827 | 0.682 | 1.003 | 0.0541 |
| Non-Hispanic White (reference) | 1 | | | | 1 | | | |
| Non-Hispanic Black | 0.555 | 0.498 | 0.618 | <.0001 | 0.664 | 0.59 | 0.748 | <.0001 |
| Other Race – Including Multi-Racial | 0.857 | 0.766 | 0.959 | 0.0073 | 1.032 | 0.835 | 1.275 | 0.7723 |
| **PIR** | | | | | | | | |
| 0-1 (reference) | 1 | | | | 1 | | | |
| 1-2 | 1.177 | 1.062 | 1.304 | 0.0018 | 1.349 | 1.181 | 1.54 | <.0001 |
| 2-3 | 1.338 | 1.195 | 1.498 | <.0001 | 1.45 | 1.251 | 1.682 | <.0001 |
| 3-4 | 1.515 | 1.344 | 1.707 | <.0001 | 1.812 | 1.547 | 2.122 | <.0001 |
| 4-5 | 1.659 | 1.456 | 1.89 | <.0001 | 1.981 | 1.669 | 2.351 | <.0001 |
| >=5 | 2.215 | 1.978 | 2.48 | <.0001 | 2.209 | 1.887 | 2.587 | <.0001 |
| **Covered by any insurance** | | | | | | | | |
| Yes | 1.338 | 1.181 | 1.516 | <.0001 | | | | |
| No (reference) | 1 | | | | | | | |
| **Covered by private insurance** | | | | | | | | |
| Yes | 1.191 | 1.072 | 1.324 | 0.0011 | 1.255 | 1.133 | 1.39 | <.0001 |
| No (reference) | 1 | | | | 1 | | | |
| **Covered by Medicare** | | | | | | | | |
| Yes | 1.217 | 1.029 | 1.439 | 0.0219 | 1.435 | 1.201 | 1.714 | <.0001 |
| No (reference) | 1 | | | | 1 | | | |
| **Covered by Medicaid** | | | | | | | | |
| Yes | 0.792 | 0.673 | 0.931 | 0.0048 | | | | |
| No (reference) | 1 | | | | | | | |
| **Covered by other government insurance** | | | | | | | | |
| Yes | | | | | 1.402 | 1.201 | 1.638 | <.0001 |
| No (reference) | | | | | 1 | | | |

**Table S3.** Reported prescription dietary supplements use by demographic characteristics among obese and control group.

| Variable | Control | | | | Obese | | | |
|---|---|---|---|---|---|---|---|---|
| | Odds ratio | 95% Wald CL | | P value | Odds ratio | 95% Wald CL | | P value |
| **Gender** | | | | | | | | |
| Male | 0.539 | 0.505 | 0.575 | <.0001 | 0.564 | 0.514 | 0.619 | <.0001 |
| Female (reference) | 1 | | | | 1 | | | |
| **Age Group** | | | | | | | | |
| 18-24 | 0.11 | 0.085 | 0.142 | <.0001 | 0.078 | 0.049 | 0.124 | <.0001 |
| 25-34 | 0.133 | 0.104 | 0.172 | <.0001 | 0.117 | 0.074 | 0.185 | <.0001 |
| 35-44 | 0.174 | 0.135 | 0.223 | <.0001 | 0.229 | 0.146 | 0.361 | <.0001 |
| 45-54 | 0.287 | 0.223 | 0.368 | <.0001 | 0.412 | 0.262 | 0.649 | 0.0001 |
| 55-64 | 0.539 | 0.418 | 0.696 | <.0001 | 0.74 | 0.468 | 1.17 | 0.198 |
| 65-74 | 0.641 | 0.509 | 0.807 | 0.0002 | 0.797 | 0.505 | 1.258 | 0.3303 |
| 75 over (reference) | 1 | | | | 1 | | | |
| **Race** | | | | | | | | |
| Mexican American | 0.479 | 0.419 | 0.547 | <.0001 | 0.442 | 0.377 | 0.518 | <.0001 |
| Other Hispanic | 0.586 | 0.501 | 0.684 | <.0001 | 0.43 | 0.349 | 0.53 | <.0001 |
| Non-Hispanic White (reference) | 1 | | | | 1 | | | |
| Non-Hispanic Black | 0.603 | 0.539 | 0.675 | <.0001 | 0.613 | 0.539 | 0.697 | <.0001 |
| Other Race – Including Multi-Racial | 0.504 | 0.447 | 0.568 | <.0001 | 0.647 | 0.513 | 0.814 | 0.0002 |
| **PIR** | | | | | | | | |
| 0-1 (reference) | 1 | | | | | | | |
| 1-2 | 0.932 | 0.835 | 1.04 | 0.2088 | | | | |
| 2-3 | 1.003 | 0.888 | 1.132 | 0.9611 | | | | |
| 3-4 | 0.921 | 0.81 | 1.047 | 0.2079 | | | | |
| 4-5 | 1.186 | 1.033 | 1.362 | 0.0153 | | | | |
| >=5 | 1.243 | 1.103 | 1.4 | 0.0003 | | | | |
| **Covered by any insurance** | | | | | | | | |
| Yes | 1.516 | 1.241 | 1.851 | <.0001 | 1.495 | 1.106 | 2.022 | 0.0089 |
| No (reference) | 1 | | | | 1 | | | |
| **Covered by private insurance** | | | | | | | | |
| Yes | 1.287 | 1.07 | 1.547 | 0.0073 | 1.57 | 1.185 | 2.08 | 0.0017 |
| No (reference) | 1 | | | | 1 | | | |
| **Covered by Medicare** | | | | | | | | |
| Yes | 2.563 | 2.079 | 3.159 | <.0001 | 3.938 | 2.886 | 5.373 | <.0001 |
| No (reference) | 1 | | | | 1 | | | |
| **Covered by Medicaid** | | | | | | | | |
| Yes | 2.016 | 1.617 | 2.514 | <.0001 | 2.5 | 1.831 | 3.415 | <.0001 |
| No (reference) | 1 | | | | 1 | | | |
| **Covered by other government insurance** | | | | | | | | |
| Yes | 1.814 | 1.479 | 2.225 | <.0001 | 2.15 | 1.596 | 2.898 | <.0001 |
| No (reference) | 1 | | | | 1 | | | |

### Performance of the Machine Learning Models for Classifying RXD and DS Use

**Table S4.** Performance of machine learning models for classifying DS use

| Model | With only demographic variables as predictors | | | | | After adding "RXD use" as a predictor | | | | |
|---|---|---|---|---|---|---|---|---|---|---|
| | **Accuracy** | **Precision** | **Recall** | **F1** | **AUROC** | **Accuracy** | **Precision** | **Recall** | **F1** | **AUROC** |
| **Logistic Regression** | **64.63** | **0.633** | **0.604** | **0.62** | **0.7** | **65.05** | **0.636** | **0.614** | **0.625** | **0.703** |
| Naïve Bayes | 64.4 | 0.621 | 0.641 | 0.63 | 0.698 | 64.1 | 0.615 | 0.647 | 0.631 | 0.699 |
| Random Forest | 63.07 | 0.612 | ,602 | ,607 | 0.67 | 62.41 | 0.602 | 0.609 | 0.606 | 0.664 |
| SMO (SVM) | 61.27 | 0.617 | 0.482 | 0.54 | 0.606 | 64.6 | 0.628 | 0.622 | 0.925 | 0.645 |

**Table S5.** Performance of machine learning models for classifying RXD use

| Model | With only demographic variables as predictors | | | | | After adding "DS use" as a predictor | | | | |
|---|---|---|---|---|---|---|---|---|---|---|
| | **Accuracy** | **Precision** | **Recall** | **F1** | **AUROC** | **Accuracy** | **Precision** | **Recall** | **F1** | **AUROC** |
| **Logistic Regression** | **74.27** | **0.77** | **0.753** | **0.76** | **0.816** | **74.3** | **0.768** | **0.758** | **0.763** | **0.818** |
| Naïve Bayes | 73.9 | 0.759 | 0.765 | 0.76 | 0.813 | 73.84 | 0.757 | 0.768 | 0.762 | 0.813 |
| Random Forest | 73 | 0.745 | 0.768 | 0.76 | 0.799 | 72.37 | 0.742 | 0.757 | 0.75 | 0.791 |
| SMO (SVM) | 71.64 | 0.749 | 0.722 | 0.74 | 0.716 | 71.64 | 0.68 | 0.71 | 0.694 | 0.716 |

**Table S6.** Performance of machine learning models for classifying DS use using PIR

| Model | With only demographic variables as predictors | | | | | After adding "RXD use" as a predictor | | | | |
|---|---|---|---|---|---|---|---|---|---|---|
| | **Accuracy** | **Precision** | **Recall** | **F1** | **AUROC** | **Accuracy** | **Precision** | **Recall** | **F1** | **AUROC** |
| **Logistic Regression** | **65.16** | **0.638** | **0.613** | **0.63** | **0.705** | **65.31** | **0.64** | **0.614** | **0.626** | **0.708** |
| Naïve Bayes | 64.65 | 0.625 | 0.634 | 0.63 | 0.701 | 64.56 | 0.62 | 0.652 | 0.636 | 0.703 |
| Random Forest | 63.97 | 0.625 | 0.6 | 0.61 | 0.689 | 63.54 | 0.617 | 0.611 | 0.614 | 0.682 |
| SMO (SVM) | 64.41 | 0.638 | 0.577 | 0.61 | 0.641 | 64.98 | 0.639 | 0.601 | 0.619 | 0.647 |

**Table S7.** Performance of machine learning models for classifying RXD use using PIR

| Model | With only demographic variables as predictors | | | | | After adding "DS use" as a predictor | | | | |
|---|---|---|---|---|---|---|---|---|---|---|
| | **Accuracy** | **Precision** | **Recall** | **F1** | **AUROC** | **Accuracy** | **Precision** | **Recall** | **F1** | **AUROC** |
| **Logistic Regression** | **74.26** | **0.772** | **0.75** | **0.76** | **0.816** | **74.27** | **0.769** | **0.756** | **0.762** | **0.818** |
| Naïve Bayes | 73.71 | 0.756 | 0.766 | 0.76 | 0.81 | 73.55 | 0.755 | 0.763 | 0.759 | 0.809 |
| Random Forest | 73.62 | 0.755 | 0.764 | 0.76 | 0.809 | 73.39 | 0.75 | 0.769 | 0.759 | 0.803 |
| SMO (SVM) | 71.64 | 0.749 | 0.722 | 0.74 | 0.716 | 71.64 | 0.749 | 0.722 | 0.735 | 0.716 |

**Table S8.** Performance of machine learning models for classifying RXD use into categories

| **Model** | Classifying into drug count groups | | | | |
|---|---|---|---|---|---|
| | **Accuracy** | **Precision** | **Recall** | **F1** | **AUROC** |
| **Logistic Regression** | **53.26** | **0.491** | **0.533** | **0.492** | **0.76** |
| Naïve Bayes | 52.69 | 0.496 | 0.527 | 0.501 | 0.755 |
| Random Forest | 50.12 | 0.473 | 0.501 | 0.482 | 0.723 |
| SMO (SVM) | 50.94 | -- | 0.509 | | 0.657 |
